# Distinct Inflammatory Profiles in Angiography-Negative Subarachnoid Hemorrhage: A Focused Case Series

**DOI:** 10.64898/2026.03.02.26347456

**Authors:** Will Remillard, Gracey Sorensen, Lauren Grychowski, David Vargas, Beatrice Hadiwidjaja, Abdelaziz Amllay, Jennifer Yan, Lena O’Keefe, Jennifer Kim, Nils Petersen, Charles Matouk, Guido J. Falcone, Kevin Sheth, Lauren H. Sansing, Jessica Magid-Bernstein

## Abstract

**Objective:** To compare early cerebrospinal fluid (CSF) cytokine profiles in intracerebral hemorrhage (ICH) versus subarachnoid hemorrhage (SAH), with a focus on angiography-negative SAH (anSAH).

**Methods:** We conducted a retrospective observational cohort study of adults with spontaneous hemorrhagic stroke (ICH or SAH). For cytokine analyses, we included patients with external ventricular drains (EVDs) and analyzed the first CSF sample obtained within 72 hours of symptom onset. Cytokines were measured using a multiplex bead-based assay and included interleukin-6 (IL-6), interleukin-8 (IL-8), vascular endothelial growth factor A (VEGF-A), C-C motif chemokine ligand-2 (CCL2), and granulocyte colony-stimulating factor (G-CSF). Cytokine concentrations were log-transformed due to non-normal distribution. Functional outcomes were assessed using the modified Rankin Scale (mRS) at discharge and 3 months.

**Results:** CSF cytokine analyses included 120 patients with available CSF samples (43 ICH and 77 SAH), while functional outcome analyses included a broader cohort of 490 patients with ICH or SAH to characterize discharge and 3-month outcomes across hemorrhage subtypes. Compared with SAH, ICH demonstrated higher early CSF log[IL-8] and log[VEGF-A] and had worse functional outcomes at discharge and 3 months. Within SAH, anSAH had higher log[IL-8] and log[VEGF-A] than aSAH, and its cytokine profile more closely aligned with that of primary ICH in hemorrhages without vascular malformations.

**Discussion:** Early CSF cytokine patterns suggest anSAH shares a more ICH-like inflammatory signature than aneurysmal SAH, supporting anSAH as a potentially biologically distinct SAH phenotype.

## INTRODUCTION

Despite advances in acute management, hemorrhagic stroke remains a highly devastating form of neurological injury. Mortality after intracerebral hemorrhage (ICH) approaches 40%, and up to 35% of patients with aneurysmal subarachnoid hemorrhage (aSAH) die, while fewer than half of survivors regain functional independence.^1–5^ The poor prognosis of both conditions highlights the need to better understand the biological mechanisms that influence recovery.

The presence of blood within the central nervous system (CNS) triggers a robust inflammatory cascade that contributes to secondary brain injury.^6–10^ In ICH, experimental models demonstrate multiple phases of inflammation: early activation of tissue-resident microglia and astrocytes, disruption of the blood-brain barrier, infiltration of circulating immune cells, and subsequent production of both pro- and anti-inflammatory molecules which aid in repair.^11^ Similarly, in aSAH, inflammatory molecules, particularly those present within the cerebrospinal fluid (CSF), have been linked to vasospasm, delayed cerebral ischemia (DCI), and worse outcomes.^12–23^

Existing studies have largely examined ICH and aSAH separately, limiting direct comparisons of inflammatory profiles across hemorrhagic stroke subtypes.^24^ Even less is known about patients with angiography-negative SAH (anSAH), a subgroup in which vascular imaging fails to identify an underlying aneurysm or vascular malformation as the etiology for the hemorrhage. AnSAH accounts for approximately 10-30% of spontaneous SAH cases, and the subgroup of anSAH patients with diffuse, non-perimesencephalic bleeding patterns in which no aneurysm is detected on initial imaging are the most diagnostically challenging, as their risk for SAH-related complications such as DCI and vasospasm remains unclear.^26^ These patients are often managed according to aSAH protocols, despite clinical uncertainty regarding whether their biology and prognosis align more closely with aneurysmal SAH or with primary ICH.^27–28^ The inflammatory profile of anSAH has not been systematically characterized, leaving an important gap in understanding.

This study seeks to directly compare CSF cytokine signatures in patients with ICH and SAH, with specific attention to the small but clinically important subset of anSAH. By examining early inflammatory mediators in CSF, we aim to (1) delineate shared and divergent immune responses between ICH and SAH, (2) highlight whether anSAH exhibits a distinct inflammatory profile, and (3) explore how these biological signatures may relate to clinical presentation, management decisions, and early outcomes.

## METHODS

### Study Design

A retrospective, observational, cohort study was performed with prior collection of biological samples and clinical data, including adult patients with spontaneous ICH or SAH admitted to the Yale Neurosciences Intensive Care Unit (NICU) between 2014 and 2025 who were enrolled in the Yale Acute Brain Injury Biorepository. Informed consent was obtained from all participants or their legally authorized representatives. CSF samples were collected from patients (n=120) in whom external ventricular drains were placed as part of their clinical care and were frozen at −80°C until cytokine analysis was performed. For the current study, only the first CSF sample collected within 72 hours of symptom onset from each patient was included to isolate early cytokine response.^11,29–30^ Patients with traumatic hemorrhage, hemorrhagic transformation of ischemic stroke, or non-vascular secondary causes of hemorrhage such as infection, tumor, or venous sinus thrombosis were excluded. For analyses of discharge outcomes and other clinical variables, the broader cohort of ICH and SAH patients including those without CSF collection (n=490) was included to capture overall clinical trends. The study was approved by the Yale University Institutional Review Board, and informed consent was obtained from patients or their legally authorized representative. Data collection and reporting adhered to STROBE guidelines for observational studies.

### Hemorrhage Classification

Hemorrhage subtype was determined based on radiographic and angiographic findings. Primary ICH was defined as intraparenchymal hemorrhage not attributable to an underlying vascular lesion, consistent with etiologies such as hypertension or cerebral amyloid angiopathy. In contrast, intraparenchymal hemorrhage resulting from rupture of an arteriovenous malformation (AVM) or dural arteriovenous fistula (dAVF) was classified as AVM-associated ICH (AVM-ICH). Patients with subarachnoid bleeding associated with a ruptured aneurysm on vascular imaging (computed tomography angiography [CTA] or digital subtraction angiography [DSA]) were categorized as aSAH. Those with spontaneous, non-traumatic SAH in which no aneurysm, AVM, or other vascular lesion was identified on initial or repeat angiography were classified as anSAH.

### Clinical and Radiographic Data Collection

Demographic variables, vascular risk factors, and hospitalization data were abstracted from the electronic medical record by a group of trained research assistants. Neuroimaging characteristics included hemorrhage location, presence of an aneurysm, ICH score, and evidence of intraventricular hemorrhage (IVH) were abstracted from the electronic medical record or calculated, as appropriate. For SAH cases, the Hunt-Hess grade and modified Fisher score were determined by review of initial imaging and documentation in the electronic medical record. The discharge and 3-month modified Rankin Scale (mRS) scores were assigned by trained raters via in person follow up visits or phone interviews, and any discrepancies in imaging characteristics or clinical variables were adjudicated during weekly consensus meetings by a group of board-certified neurointensivists (JMB, LO, JK, NP). Reviewers were blinded to cytokine data.

### CSF Sampling and Cytokine Analysis

CSF samples were collected between presentation and 72 hours after symptom onset, immediately centrifuged to remove cellular debris, aliquoted, and stored at −80 °C until analysis. Cytokine concentrations were quantified using a multiplex bead-based assay (cytometric bead array; BD Biosciences, Franklin Lakes, NJ) according to manufacturer protocols. The personalized cytokine and chemokine panel included interleukin-6 (IL-6), interleukin-8 (IL-8), vascular endothelial growth factor A (VEGF-A), C-C motif chemokine ligand-2 (CCL2), and granulocyte colony-stimulating factor (G-CSF). The lower limit of detection (LLD) corresponded to the concentration of the lowest standard for each analyte, and any values below the LLD were set at half of the LLD for each experiment.

### Statistical Analysis

Cytokine concentrations were non-normally distributed, and as such were log transformed for all analyses. Linear regression models were used to evaluate the association between log-transformed cytokine levels and (1) hemorrhage location (ICH vs SAH) and (2) vascular etiology (presence vs absence of vascular malformation). For primary analyses, hemorrhage subtype was compared as ICH with SAH, with ICH including both primary ICH and AVM-ICH and SAH analyzed as a single group including both aSAH and anSAH. This approach reflects that early clinical management is largely standardized across spontaneous SAH phenotypes, with the major distinction being aneurysm securement in patients found to have an aneurysm. Secondary analyses then examined more specific etiologic comparisons using the following pairwise groups: aSAH with anSAH, anSAH with primary ICH, aSAH with AVM-ICH, primary ICH with AVM-ICH, and AVM-ICH with anSAH. Comparisons of discharge and 3-month mRS scores dichotomized into favorable (mRS 0-4) and unfavorable (mRS 5-6) outcomes between hemorrhage subgroups, were performed using chi-square testing. Statistical analyses were performed in R (version 4.5.1), and significance was defined as p < 0.05.

## RESULTS

CSF samples were collected from 43 patients with ICH (36 primary, 7 AVM-ICH) and 77 patients with SAH (72 aSAH, 5 anSAH). Compared with SAH overall, ICH patients demonstrated higher CSF log[IL-8] (β = 0.42, 95% CI 0.13–0.74, p = 0.004) and log[VEGF-A] (β = 0.81, 95% CI 0.47– 1.21, p < 0.001; Fig. 1c). Within the SAH group, anSAH patients had higher CSF log[IL-8] (β = −0.85, 95% CI −1.75– −0.11, p = 0.030) and log[VEGF-A] (β = −1.18, 95% CI −2.29– −0.35, p = 0.003) compared to aSAH patients (Fig. 1c). Among hemorrhages with vascular etiologies, aSAH differed from AVM-ICH for CSF log[VEGF-A] (β = −0.94, 95% CI −1.77– −0.26, p = 0.011; Fig. 1c), but there were no differences in any of the other measured cytokines.

**Figure 1.**
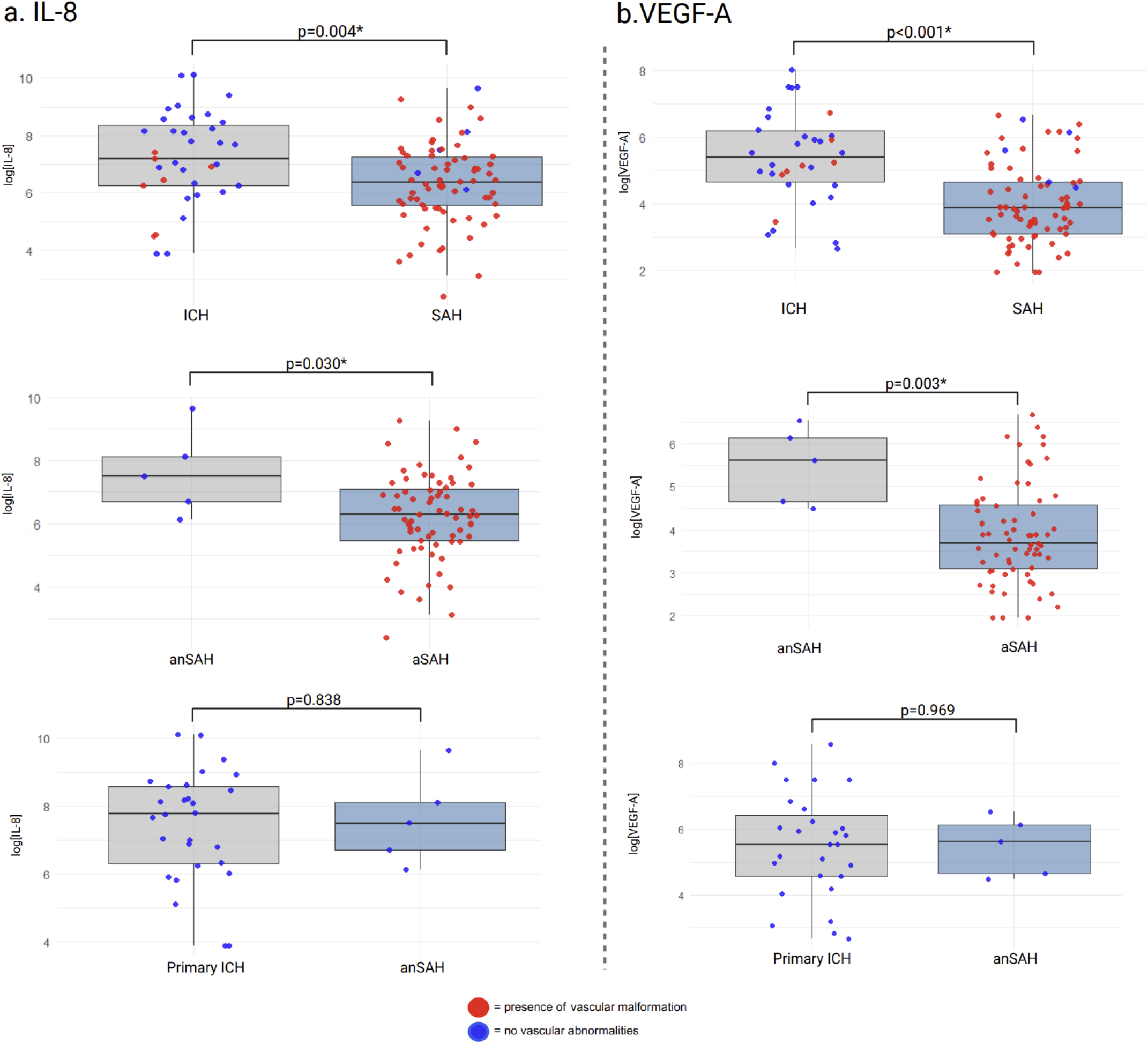
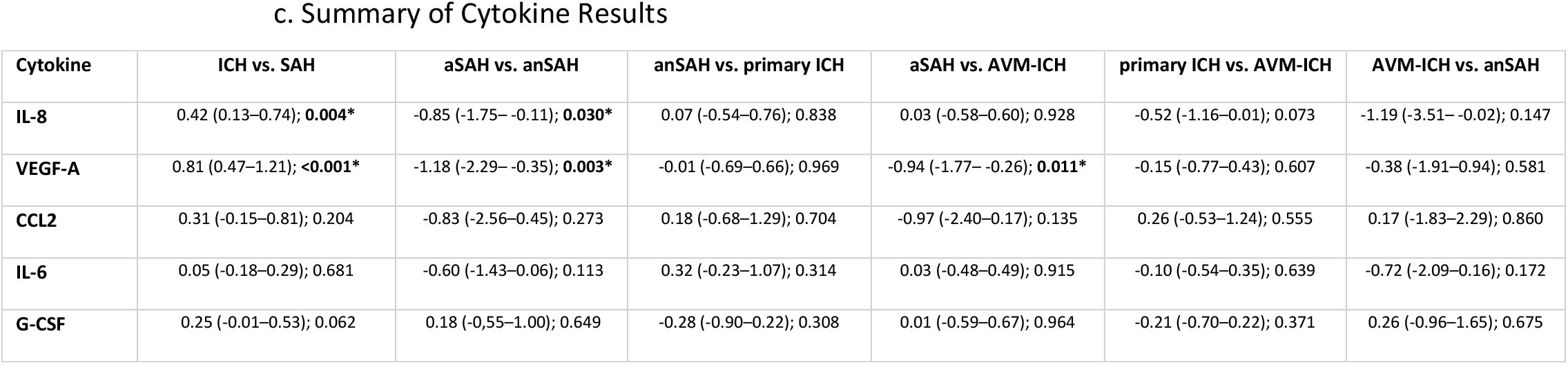
Cytokine Levels vs. Hemorrhage Type. Boxplots of CSF log[IL-8] (a) and log[VEGF-A] (b) collected on days 0–3 show higher cytokine levels in ICH vs SAH and in anSAH vs aSAH, with no significant differences between anSAH and primary ICH. P values from linear regression models of log-transformed cytokine levels are shown above each comparison. Panel (c) summarizes regression outputs for all cytokines across hemorrhage subgroups, reported estimates with 95% confidence intervals and p values (beta (95% CI); p-value). Cerebrospinal fluid, CSF; interleukin-8, IL-8; vascular endothelial growth factor A, VEGF-A; intracerebral hemorrhage, ICH; angiography-negative subarachnoid hemorrhage, anSAH; aneurysmal subarachnoid hemorrhage, aSAH; beta, β; confidence interval, CI.

Additionally, in the larger cohort of 490 patients with outcome data collected, ICH hemorrhage location was associated with worse functional outcomes at both discharge and 3 months, when compared with SAH (OR = 3.31, 95% CI 2.09–5.28, p < 0.001 and OR = 2.04, 95% CI 1.30–3.22, p = 0.045, respectively; Fig. 2a). Among all SAH patients, higher CSF log[G-CSF] was associated with poor functional outcome (OR = 1.41, 95% CI 1.01–1.98, p = 0.046; Fig. 2b).

**Figure 2.**
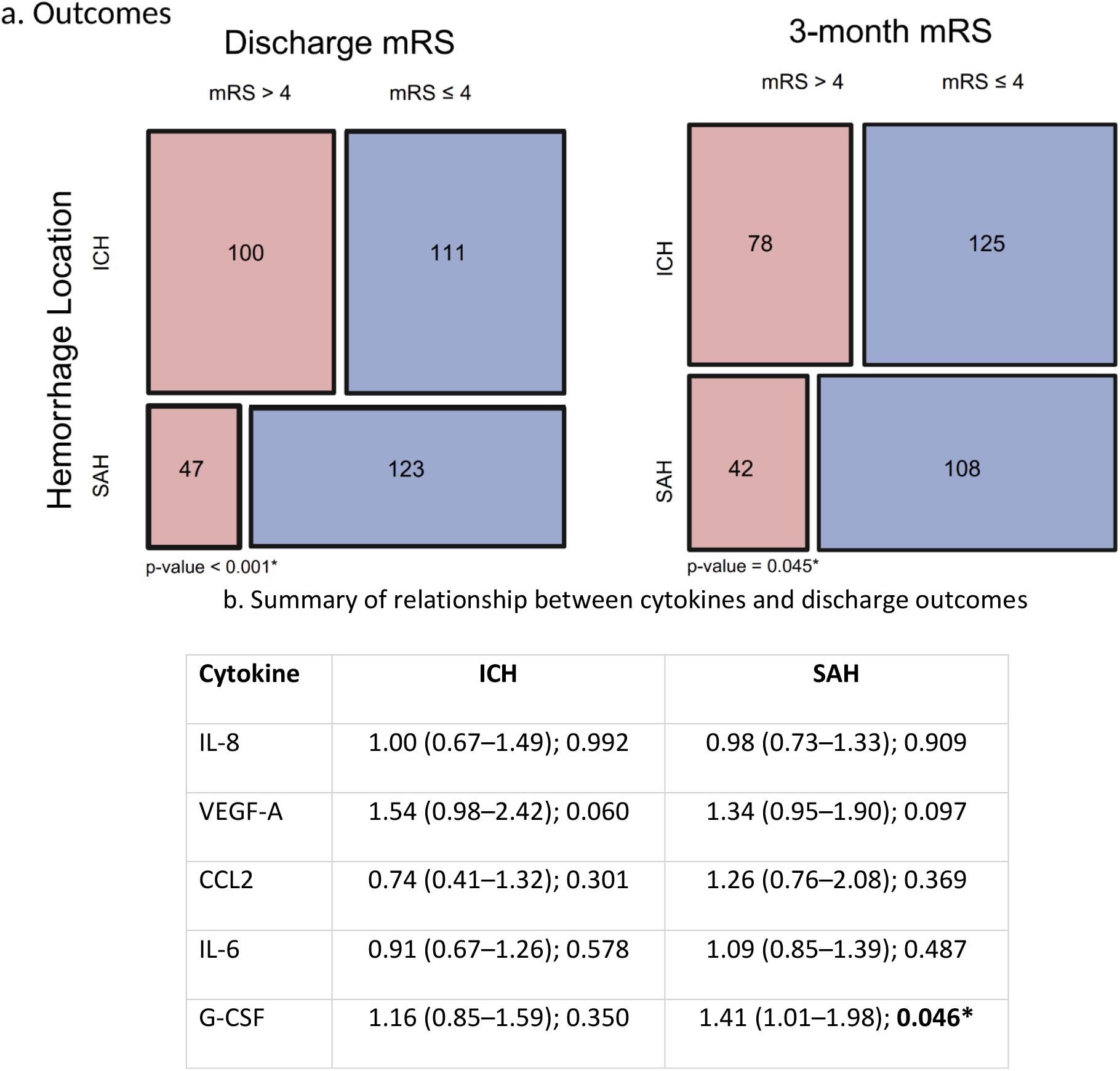
Outcome Analysis between Hemorrhage Location and CSF Cytokine. Mosaic plots of discharge and 3-month mRS by hemorrhage location (ICH vs SAH) show a higher proportion of poor outcome (mRS > 4) in ICH at both time points (chi-squared p<0.001 at discharge, p=0.045 at 3 months). Rectangle area is proportional to group counts and color reflects outcome (red = mRS > 4, blue = mRS ≤ 4). Panel (b) shows cytokine–outcome regression results for ICH and SAH. Among SAH patients, only higher CSF log[G-CSF] demonstrated a weak association with poor functional outcome ([OR] 1.41 [95% CI] 1.01–1.98], p=0.046). Cerebrospinal fluid, CSF; interleukin-8, IL-8; vascular endothelial growth factor A, VEGF-A; intracerebral hemorrhage, ICH; subarachnoid hemorrhage, SAH; odds ratio, OR; confidence interval, CI, modified Rankin score, mRS.

When directly comparing the two hemorrhage subtypes without vascular malformations, CSF cytokine concentrations did not differ between anSAH and primary ICH for any of the measured cytokines (all p>0.05; Fig. 1c).

## DISCUSSION

In this study comparing early CSF cytokine profiles across hemorrhagic stroke subtypes, we found that IL-8 and VEGF-A were higher in ICH than in SAH, and that anSAH shared an inflammatory signature more similar to ICH than to aSAH. We did not find a difference in CSF concentrations of the other measured cytokines, including IL-6, CCL2, and G-CSF, between SAH and ICH samples or between samples collected from patients with or without a vascular etiology for their hemorrhage. These findings align with prior evidence that pro-inflammatory cytokines are elevated after hemorrhagic stroke.^30–33^ In the current study, we observed higher CSF IL-8 and VEGF-A following anSAH and ICH when compared to aSAH. While these results are preliminary and hypothesis generating given the small sample size, it suggests that blood contact with the parenchyma or ventricular surfaces, rather than aneurysmal rupture, drives early endothelial cell production of VEGF-A and neutrophil recruitment by IL-8. In contrast to the similarities we observed between ICH and anSAH, AVM-ICH does not simply mirror aSAH, and our pairwise comparisons suggest differences between these vascular etiologies, particularly for VEGF-A. Together, these patterns support the interpretation that anSAH shares an early CSF inflammatory signature more similar to ICH than to aSAH, while also suggesting that the presence of a vascular lesion is not sufficient to predict cytokine profile.

IL-8, a potent neutrophil chemoattractant, mediates early innate immune activation and amplifies secondary injury via oxidative stress, additional cytokine release, and blood-brain barrier disruption. High CSF IL-8 therefore implies robust local immune cell recruitment to sites of parenchymal or ventricular blood exposure, processes pronounced in ICH and, as our findings suggest, in anSAH.^34^ VEGF-A, a key regulator of vascular permeability and endothelial survival, promotes blood-brain barrier leakage and vasogenic edema in the acute phase. Higher CSF VEGF-A in ICH and anSAH may reflect greater endothelial activation and blood-brain barrier dysfunction, consistent with a stronger early blood-CSF inflammatory stimulus in these groups compared with aSAH.^35^ Interestingly, CSF VEGF-A concentrations were higher in hemorrhages without an underlying vascular malformation than in aneurysmal or AVM-related bleeds, despite the classical association between VEGF-A and angiogenesis.^35^ The reason for this unexpected finding remains unclear, but it suggests that primary parenchymal or periventricular bleeding may inflict more extensive endothelial injury and hypoxia-driven VEGF-A release than rupture of a pre-existing malformation, or perhaps that VEGF-A production is intrinsically suppressed in the setting of an underlying vascular malformation. Collectively, these data support the concept that anSAH represents a distinct biological entity within the spectrum of spontaneous SAH, with an inflammatory profile that tracks more closely with ICH than with aSAH. These findings warrant further studies and may have implications for further immune-targeted treatments for ICH and SAH.

This study is limited by a small sample size, single-center design, and CSF collection confined to an early window (≤72 hours). Functional outcome was measured by discharge and 3-month mRS, capturing early disability but not long-term recovery.

This study is meant to be hypothesis generating, as we report a novel analysis of CSF cytokines in different hemorrhage subtypes. Validation in larger, multicenter cohorts and mechanistic studies of IL-8 and VEGF-A signaling may clarify the role that these pathways may play in secondary injury and outcome after hemorrhagic stroke. Ultimately, integrating inflammatory profiling into practice could enable identification of biomarkers to guide immune-targeted medications, informing management across the hemorrhagic stroke spectrum.

## Data Availability

All data produced in the present study are available upon reasonable request to the authors.

## ACKNOWLEDGEMENTS

Figures were created with BioRender.com

## TABLES and FIGURES

**Table 1.**
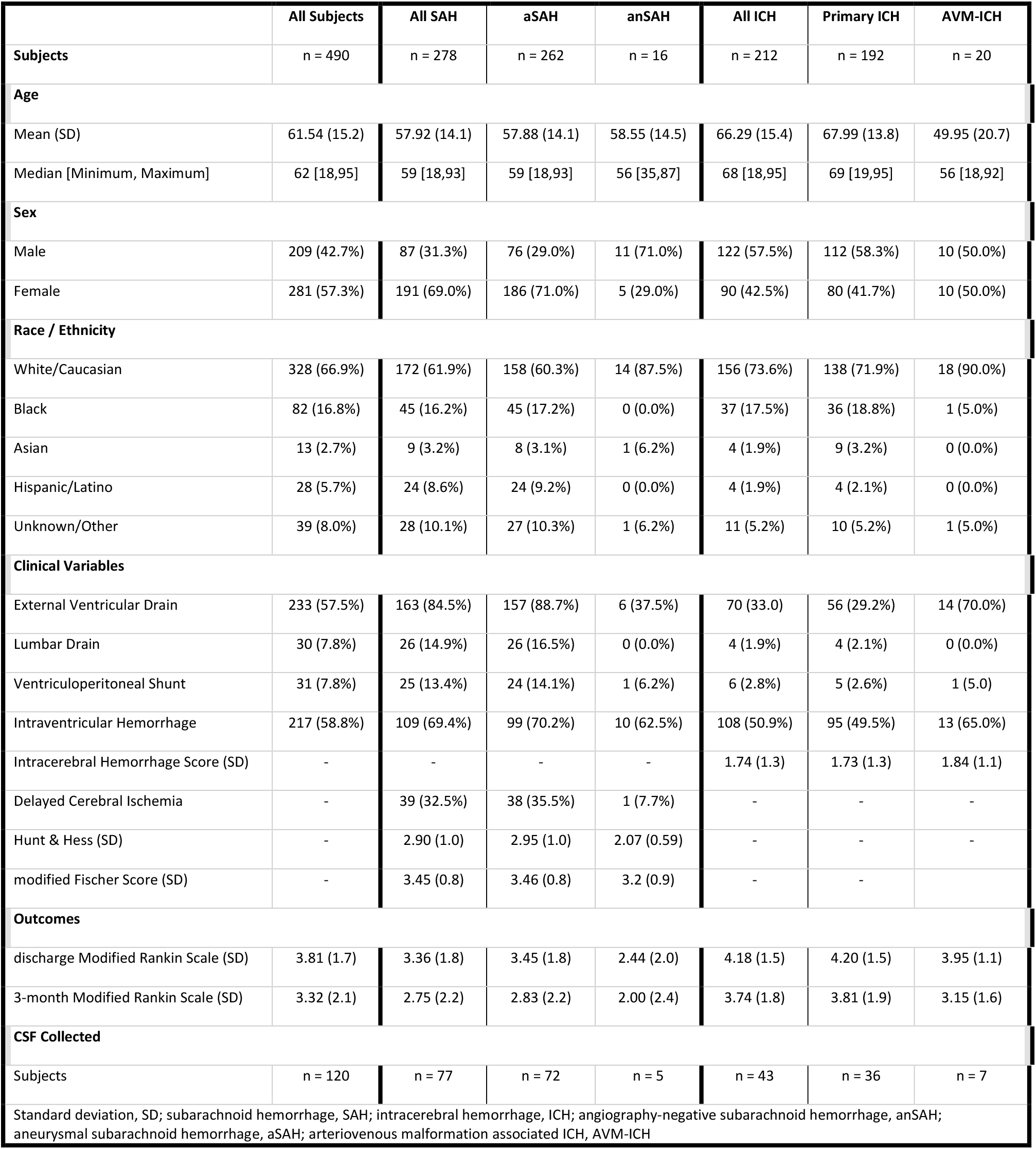
Demographics.

